# Absolute and Relative Effectiveness of High-Dose vs Standard-Dose Influenza Vaccines Against Severe Outcomes in Older Adults

**DOI:** 10.64898/2026.10.05.26364731

**Authors:** Stéphane Le Vu, Nicolás Gutiérrez-Melo, Thomas Cartailler, Marie-Joelle Jabagi, Jérémie Botton, Valérie Olié, Alain Weill, Rosemary Dray-Spira, Mahmoud Zureik

## Abstract

**Importance:** Several influenza vaccine products are available for older adults, but evidence combining both their absolute effectiveness and head-to-head comparisons within the same large population remains scarce, limiting product-specific vaccination guidance for this high-risk group.

**Objective:** To estimate the relative and absolute effectiveness of influenza vaccines against influenza-related hospitalization and in-hospital mortality among older adults.

**Design, Setting, and Participants:** Nationwide cohort study using the French National Health Data System (SNDS), including all adults born in 1958 or earlier, living in metropolitan France, at the start of the 2023-2024 campaign. Relative effectiveness (rVE) was assessed using a landmark design with exposure fixed before epidemic onset. Absolute effectiveness (VE) was assessed using a sequential trial emulation with weekly matching of newly vaccinated individuals to yet unvaccinated controls. Follow-up ran from December 5, 2023 to April 19, 2024.

**Exposures:** Vaccination with a high-dose (Efluelda) or one of three standard-dose quadrivalent vaccines (Influvac, Fluarix, Vaxigrip), compared individually or by dosage category, and against no vaccination.

**Main Outcomes and Measures:** Hospitalization with a diagnosis of influenza and death during such hospitalization. Effectiveness was estimated as 1 minus the hazard ratio from weighted Cox models, with corresponding numbers needed to vaccinate to prevent one case.

**Results:** Among 13.8 million eligible individuals, 5.7 million (41%) were vaccinated at the beginning of follow-up. The high-dose vaccine showed greater rVE than standard-dose vaccines against hospitalization (27% [95%CI, 23 to 31%]) and in-hospital death (27% [95%CI,14 to 38%]). Absolute VE was 39% (95%CI, 38 to 41%) against hospitalization and 45% (95%CI, 40 to 49%) against in-hospital death, and was higher for the high-dose vaccine (50%) than standard-dose vaccines (36%) against hospitalization. This difference was most evident among adults 70 years and older, while overall effectiveness declined with age.

**Conclusions and Relevance:** In this study, influenza vaccination conferred moderate protection against severe outcomes in older adults, and high-dose vaccination was associated with significantly greater effectiveness than standard-dose vaccines, particularly among adults 70 years and older. These findings support product-level differentiation in vaccination strategy for this population.

**Key points:** *Question:* What is the absolute effectiveness of influenza vaccination and the relative effectiveness of high-dose versus standard-dose vaccines against severe influenza outcomes in older adults?

*Findings:* In this nationwide cohort study of 13.8 million older adults in France, vaccination was associated with 39% lower risk of influenza-related hospitalization and 45% lower risk of death during those hospitalizations compared with no vaccination. The high-dose vaccine was associated with greater absolute effectiveness than standard-dose vaccines against hospitalization (50% vs 36%), a difference most evident among adults 70 years and older, and with significantly greater relative effectiveness against hospitalization (27%) as well.

*Meaning:* These findings suggest that high-dose influenza vaccination provides added protection over standard-dose formulations in older adults, particularly from age 70 onward, and may support product-specific vaccination recommendations for this population.

## Introduction

Routine annual vaccination is recommended in many countries for people 65 years and older, in order to prevent the severe consequences of seasonal influenza such as hospitalization and death^1^. Several vaccines are commonly available across health systems, including newer enhanced vaccine formulations developed to specifically address the decline in immune response for this population.

Comparative effectiveness between products have been assessed in randomized controlled trials and observational studies. High-dose or adjuvanted vaccines, for example, may offer superior protection over standard-dose formulations^2–5^. One study in Nordic countries reported brand-specific absolute effectiveness, but could not draw head-to-head comparisons between vaccines, as recipients of respective products differed systematically by age^6^.

In France several products are licensed for people 65 and over, but with no preferential age-based indication within this group. The product choice is thus left to the discretion of the prescriber or pharmacist. In particular during the season 2023-2024, a high-dose vaccine was used alongside three other quadrivalent standard-dose vaccines. This flexibility in the distribution provides an opportunity for relatively balanced vaccine usage and to measure head-to-head comparison between products across all age brackets. As health authorities and practitioners increasingly seek to differentiate between available vaccine options, particularly for high-risk groups such as older adults, there is a clear need for observational studies assessing the effectiveness of individual vaccine products against severe outcomes.

Here, we aimed to estimate at a nationwide scale both the relative (head-to-head comparisons between vaccine products) and absolute effectiveness (each product compared with no vaccination) of vaccines in preventing influenza-related hospitalization and in-hospital mortality among elderly persons.

## Methods

### Data source

We analyzed data from the National Health Data System (Système National des Données de Santé - SNDS), which contains reimbursement data for healthcare expenditure for almost all people living in France^7^ (Details in eMethods).

### Study population

We included all adults registered to the SNDS, born in 1958 or earlier, residing in metropolitan France, and who had received at least one healthcare service or treatment in the 12 months prior to the start of the vaccination campaign on October 17, 2023. This last criterion was intended to rule out individuals who were not currently living in France, for whom vaccination or potential outcomes would not be observable. As per the age-based recommendations, all these individuals were eligible to receive a flu vaccine free of charge. We excluded individuals residing in nursing homes with an on-site pharmacy, whose medication costs are reimbursed through a lump-sum allocation, for whom vaccine administration cannot be tracked on an individual basis. These individuals account for approximately 20% of all nursing home residents.

### Outcomes

The two primary outcomes were (i) hospitalization with a diagnosis of influenza (primary, related, or associated diagnosis) and (ii) death from any cause during such hospitalizations (see eTable *1* for corresponding ICD-10 codes). A set of secondary outcomes was defined to assess the potential indirect effect of protection against severe influenza and comprised hospitalization with the following diagnoses: (i) pneumonia, (ii) any cardiac disorder, and (iii) in-hospital death following a hospitalization with any diagnosis. We also evaluated two hospital diagnoses which were supposed to be unrelated to vaccine exposure and the risk of influenza as negative control outcomes : (i) traumatic injury and (ii) cataract. For all outcomes, only the first hospitalization during follow-up was considered.

### Study designs

We devised a specific cohort study design for each objective of estimating relative (rVE) and absolute vaccine effectiveness (VE). To estimate comparative effectiveness between vaccine products, we used a landmark design where vaccine exposure is fixed at a time zero, before the epidemic starts. For the derivation of absolute effectiveness, we emulated a sequence of target trials to account for a time-varying exposure. Enrollment began on December 5, 2023 (time zero), i.e. 14 days before the epidemic officially started^8^, and ended on April 19, 2024, 60 days past the end of the epidemic. The 14-day period was set in line with the lag between vaccination and the building of a full immunity. Individuals were classified as exposed from 14 days after vaccination. Until then, they were classified as unexposed, along with individuals who had not been vaccinated. This ensured that all persons already vaccinated at the first day of enrollment could be considered exposed at the start of the epidemic. Vaccine exposure was considered by individual product or by category of products differentiating standard-dose (SD) and high-dose (HD) vaccines. Even though all vaccines used in the season were quadrivalent, for the sake of brevity, we use their brand names without the “Tetra” suffix throughout the text.

### Landmark analysis

In this design, exposure status for each vaccine was defined at enrollement - by which time 89% of all individuals vaccinated during the season had already been vaccinated - and as in an intention-to-treat analysis is considered fixed. We matched recipients of the vaccine of interest and of the comparator vaccine on baseline characteristics with a constraint of exact matching on sex, age class, administrative region of residence and nursing home residency. Beside these 4 matching variables, baseline covariates were variables potentially associated with a risk of severe influenza or with vaccine exposure: number of primary care contacts within 12 months, affiliation to complementary solidarity health insurance, and history of comorbidities within 5 years (see eMethods). We used the generalized full matching method and derived corresponding matching weights^9^.

### Sequential trial analysis

We emulated a sequential trial of the effect of vaccine exposure compared to no vaccination. Each week of the study period, a risk set was defined by matching all newly exposed persons with yet unexposed controls, with a 1:1 ratio. We used exact matching on the variables sex, age class, region and nursing home residency. To further balance baseline confounders between vaccinated and unvaccinated, we calculated a propensity score on the above mentioned baseline characteristics. A distinct propensity score model was fitted for each risk set. The first (and largest) risk set thus comprised all exposed individuals in the pre-epidemic period, along with their matched unexposed controls. The same unexposed subjects could be sampled multiple times at different periods, or could eventually become exposed later in the season and considered as such in other risk sets. For this reason, and because the exposed and unexposed groups are defined differently, the sample sizes and numbers of events cannot be compared between the landmark analysis and this sequential trial analysis where exposure is time-dependent. The start of follow-up was reset to zero in each risk sets and data from all risk sets were pooled together for further analyses.

### Statistical Analysis

We calculated average effect of treatment among the treated (ATT) weights from either the generalized full matching or the propensity score procedures in the two designs outlined above. We derived Kaplan-Meier estimates of cumulative incidence by exposure groups and hazard ratios from weighted cox models. Follow-up was censored at the earliest of the outcome of interest, death, or end of the study period. For the sequential design, standard errors of weighted Cox coefficients were estimated using a robust sandwich (Huber-White) variance estimator, with score residuals clustered by weekly matched risk set. A sensitivity analysis was conducted by assessing the clustering at subject level, and using a two-way cluster-robust sandwich variance estimator to assess whether within-subject correlation across weekly risk sets materially affected inference^10^. VE was estimated as one minus the hazard ratio. It represents the proportion of events prevented by vaccination in the vaccinated population, or for rVE, the proportion of events prevented by using the vaccine of interest instead of the comparator. We calculated the number of persons needed to vaccinate (NNV) to avert one event by dividing the number persons exposed by the number of events prevented and expressed their 95% confidence interval following Altman^11^ (see eMethods). The analyses were stratified into 4 age classes (64-69, 70-74, 75-79, 80 and older), which were determined by quartiles of the age distribution.

## Results

After excluding 96,808 (0.7%) persons residing in nursing homes with an on-site pharmacy, a total of 13,788,297 persons were included and followed-up from December 5, 2023 to April 19, 2024. Their baseline characteristics by vaccination status are shown in Table 1. At the beginning of the follow-up period, 5,689,177 (41%) persons had received an influenza vaccine for at least 14 days (SD: Influvac 41 %, Vaxigrip 21 %, Fluarix 6.8 %, and HD: Efluelda 32 %). The distribution across age groups of vaccine recipients during the whole season is shown in eTable *2* and indicates that each product was used by people of all ages. Vaccine recipients were older, more often male, nursing home residents and had a higher prevalence of comorbidity and frequency of healthcare contacts (Table 1).

**Table 1:**
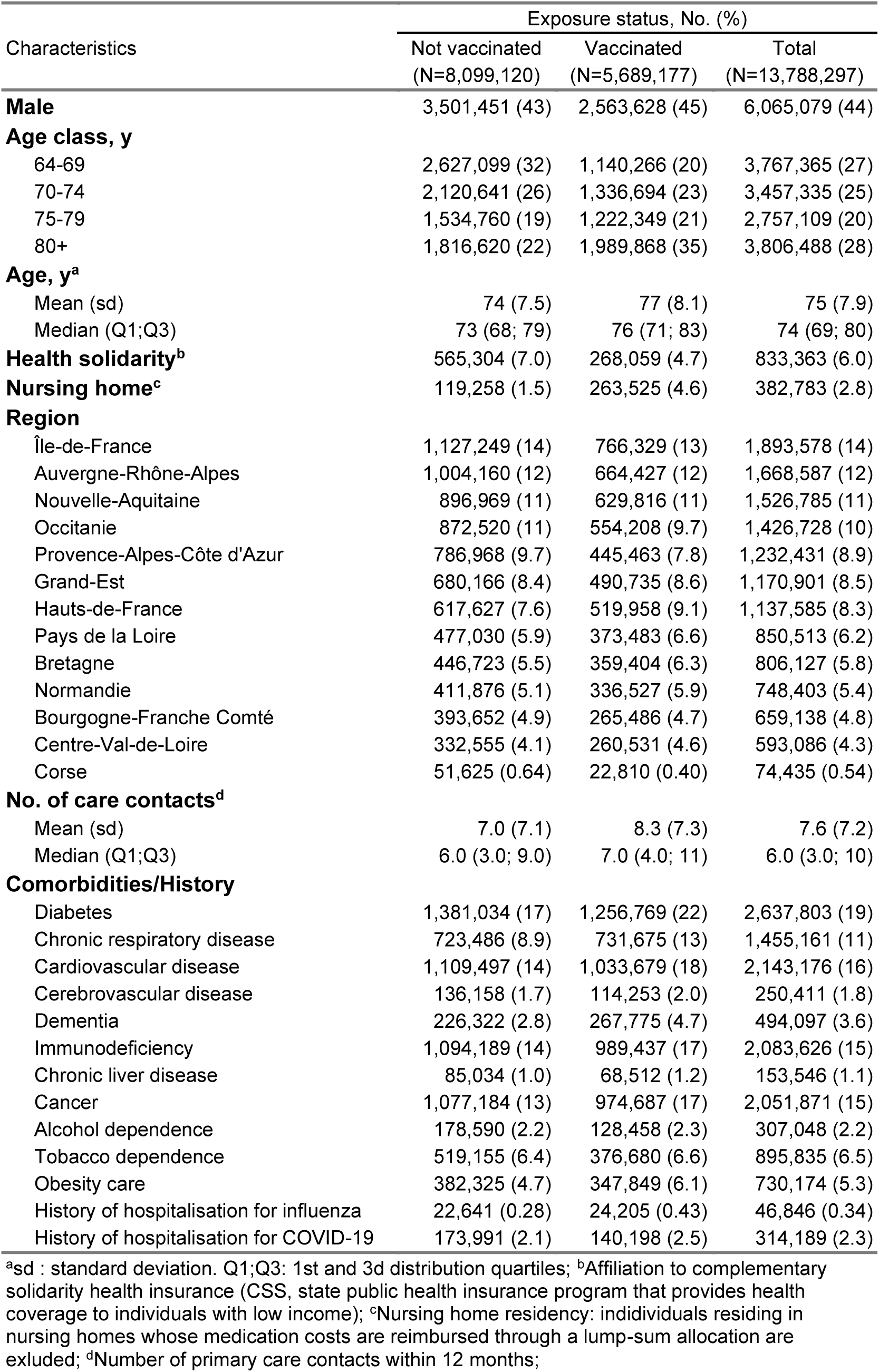
Characteristics of study population by vaccination status at baseline.

At the end of the study period, a total of 25,214 hospitalizations with a diagnosis of influenza and 2,199 deaths during these hospitalizations had been recorded.

Relative effectiveness

eTable *3* compares the characteristics of persons having received a SD vs a HD vaccine. Vaccinated persons were more similar to one another across the two groups than they were to the unvaccinated population. However, HD vaccine users were even older, more often female, nursing home residents, and had a higher prevalence of comorbidity (except tobacco dependence) and frequency of healthcare contacts than standard vaccines users. All variables were well balanced between the study groups after matching (eFigure *1*). Among these vaccinated persons at baseline, 10,296 hospitalizations for influenza were documented and 932 resulted in death, over a total follow-up of 136 days. Median time from baseline to hospitalization for influenza was 57 days (quartiles 41 and 70 days), and from baseline to death during those hospitalizations was 67 days (quartiles 51 and 82 days). Figure 1(A and B) shows the cumulative incidence curves for those two primary outcomes. For both influenza-related hospitalization and death during those hospitalizations, incidence was lower in HD vaccine (Efluelda) recipients. Estimates of rVE against hospitalization for influenza and death following such hospitalization are presented in Table 2 and eTable *4*. The HD vaccine offers greater overall protection – with an rVE of 27% (95%CI, 23% to 31%) for hospitalization for influenza and 27% (95%CI, 14% to 38%) for in-hospital death – in the overall comparison against all SD vaccines. These rVE estimates translate into NNV with the HD instead of SD vaccines between 1,400 and 2,000 to prevent one influenza-related hospitalization, and between 9,600 and 37,000 to prevent one death following such influenza hospitalization. In head-to-head comparisons of individual vaccines, and for both hospitalization (Table 2) and death (eTable *4*), Efluelda vaccine has a greater effectiveness than Influvac and Vaxigrip vaccines but the difference is not observed when compared to Fluarix vaccine. eFigures *2* and *3* show the cumulative incidence curves in HD and SD vaccine recipients for the secondary (pneumonia, any cardiac disorder, and all cause in-hospital death) and negative outcomes (traumatic injury and cataract). The corresponding estimates of rVE are shown in eTable *5*. The HD vaccine was marginally more effective at preventing hospitalization for a cardiac disorder (1.6% [95%CI, 0.70% to 2.6%]) but no difference in effectiveness is observed between vaccine types for pneumonia, all-cause in-hospital death or for traumatic injury. Unexpectedly, we also observed a slightly reduced risk of cataract hospitalization in HD vaccine recipients. Figure 2(A) and eTable *6* show the estimated rVE between HD and SD vaccines for hospitalization for influenza by age class.

**Figure 1:**
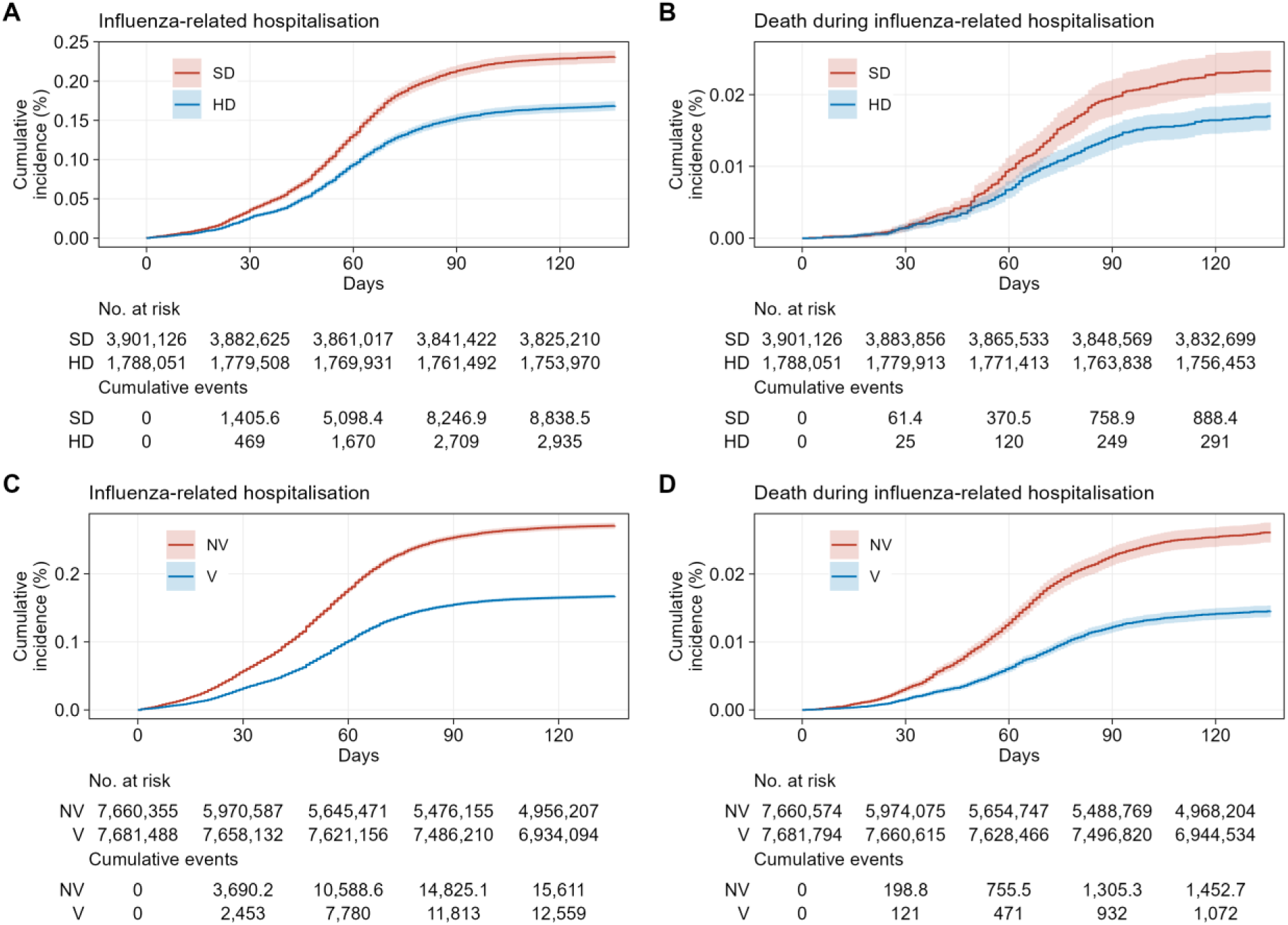
Cumulative incidence curves for primary outcomes. Panels **A** and **B** are results of the landmark analysis of relative effectiveness. Panels **C** and **D** are results of the sequential trial analysis of absolute effectiveness. Cumulative incidence curves over the 137-day follow-up were obtained from ATT-weighted Kaplan-Meier estimates, with weights derived from generalized full matching in the landmark analysis and from propensity score weighting in the sequential trial emulation. Abbreviations: SD Standard-dose vaccines; HD High-dose vaccine; NV Not vaccinated; V Vaccinated.

**Figure 2:**
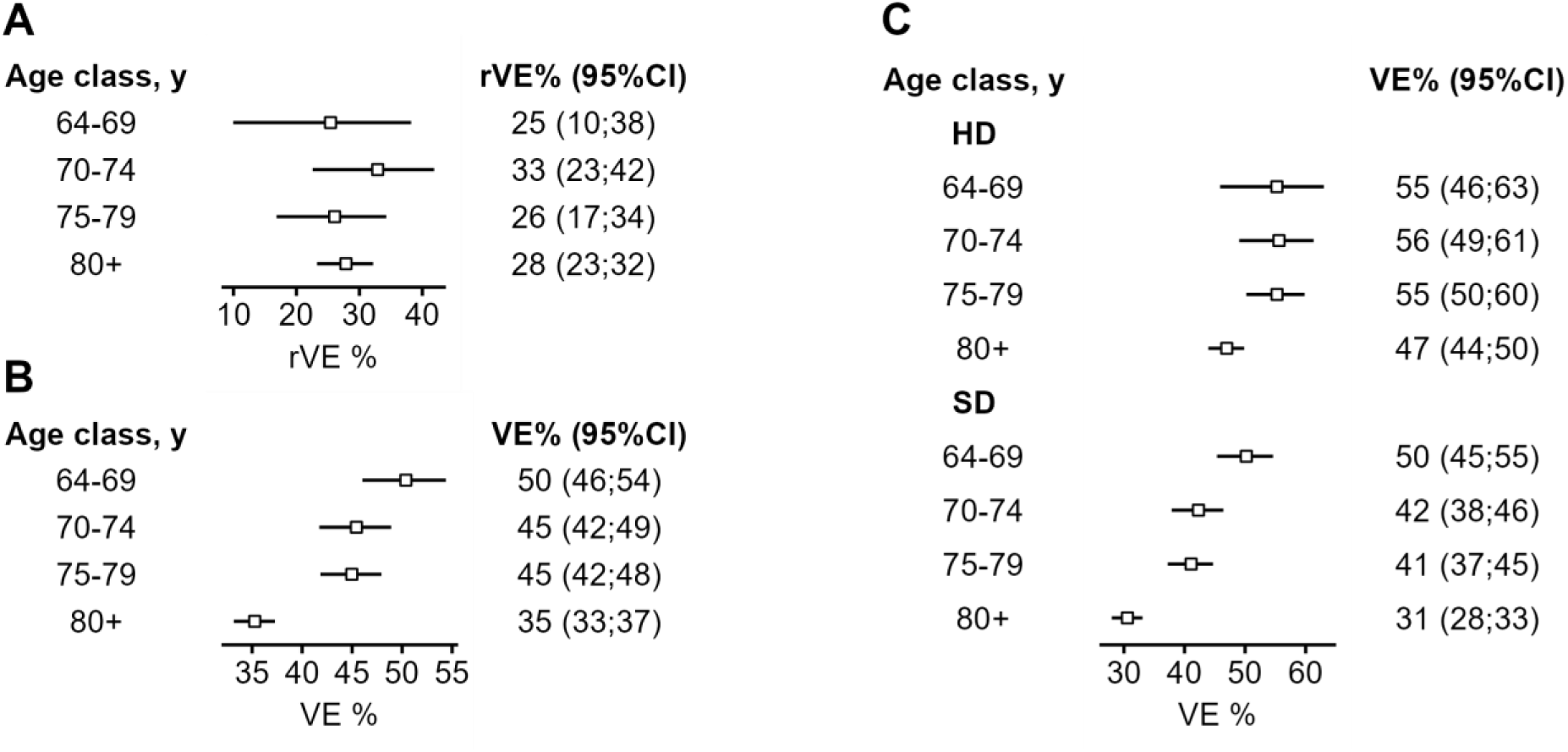
Vaccine effectiveness against influenza-related hospitalization by age class. **A**: Relative vaccine effectiveness (rVE) of high-dose (HD) vs standard-dose (SD) vaccines on hospitalization for influenza, by age class. **B**: Overall absolute effectiveness of vaccination against influenza-related hospitalization by age class. **C**: Absolute vaccine effectiveness (VE) against influenza-related hospitalization of high-dose (HD) and standard-dose (SD) vaccines compared to no vaccination, by age class.

**Table 2:** Relative vaccine effectiveness against influenza-related hospitalization.

| Reference exposure |  |  | Exposure of interest |  |  | rVE<br>% [95%CI]<sup>a</sup> | NNV<br>No. [95%CI]<sup>b</sup> |
| --- | --- | --- | --- | --- | --- | --- | --- |
| Vaccine | Exposed<br>No. | Events<br>No. (%) | Vaccine | Exposed<br>No. | Events<br>No. (%) |  |  |
| SD<sup>c</sup> | 3,901,126 | 7,309 (0.19) | HD<sup>d</sup> | 1,788,051 | 2,987 (0.17) | 27 [23; 31] | 1,600 [1,400; 2,000] |
| Fluarix | 389,770 | 614 (0.16) | Efluelda | 1,788,051 | 2,987 (0.17) | 4.8 [-8.1; 16] | 12,000 [H 8,000; ∞; B 3,100] |
| Influvac | 2,327,657 | 4,544 (0.20) | Efluelda | 1,788,051 | 2,987 (0.17) | 27 [23; 31] | 1,600 [1,300; 2,000] |
| Vaxigrip | 1,183,674 | 2,151 (0.18) | Efluelda | 1,788,051 | 2,987 (0.17) | 23 [17; 28] | 2,000 [1,500; 2,900] |
| Influvac | 2,327,657 | 4,544 (0.20) | Fluarix | 389,770 | 614 (0.16) | 25 [18; 31] | 1,900 [1,400; 3,000] |
| Vaxigrip | 1,183,674 | 2,151 (0.18) | Fluarix | 389,770 | 614 (0.16) | 20 [12; 28] | 2,500 [1,700; 4,800] |
| Influvac | 2,327,657 | 4,544 (0.20) | Vaxigrip | 1,183,674 | 2,151 (0.18) | 0.67 [-5.3; 6.3] | 81,000 [H 11,000; ∞; B 8,200] |

<sup>a</sup>Relative vaccine effectiveness; <sup>b</sup>Number needed to vaccinate. When vaccine effectiveness is negative, indicating a higher risk of the outcome with the exposure of interest than the reference, the negative NNV corresponds to the number needed to harm (NNVH). For intervals encompassing infinity, see the methods section; <sup>c</sup>Any standard-dose vaccine; <sup>d</sup>High-dose vaccine (here Efluelda);

Point estimates of rVE are between 25 and 33% with no clear age tendency and confidence intervals largely overlap. However, the smallest NNV (or greatest number of hospitalizations averted by using a HD instead of a SD vaccine) is estimated in the older age class of 80 years and over (eTable *6*).

### Absolute effectiveness

In the sequence of 19 weekly trials, a total number of 7.9 million individuals were vaccinated throughout the season, of which 7.7 million (97%) were matched to an unvaccinated control (eTable *7*). Figure 1(C and D) shows the cumulative incidence curves in unvaccinated and vaccinated groups for the two primary outcomes and indicates a lower incidence in vaccine recipients for both influenza specific outcomes. The overall absolute vaccine efficacy (VE) is estimated at 39% (95%CI, 38% to 41%) against hospitalization and 45% (95%CI, 40% to 49%) against death during influenza-related hospitalization (Table 3 and eTable *8*). When the analysis is broken down by vaccine type, the VE of the HD vaccine is higher than that of the

**Table 3:**
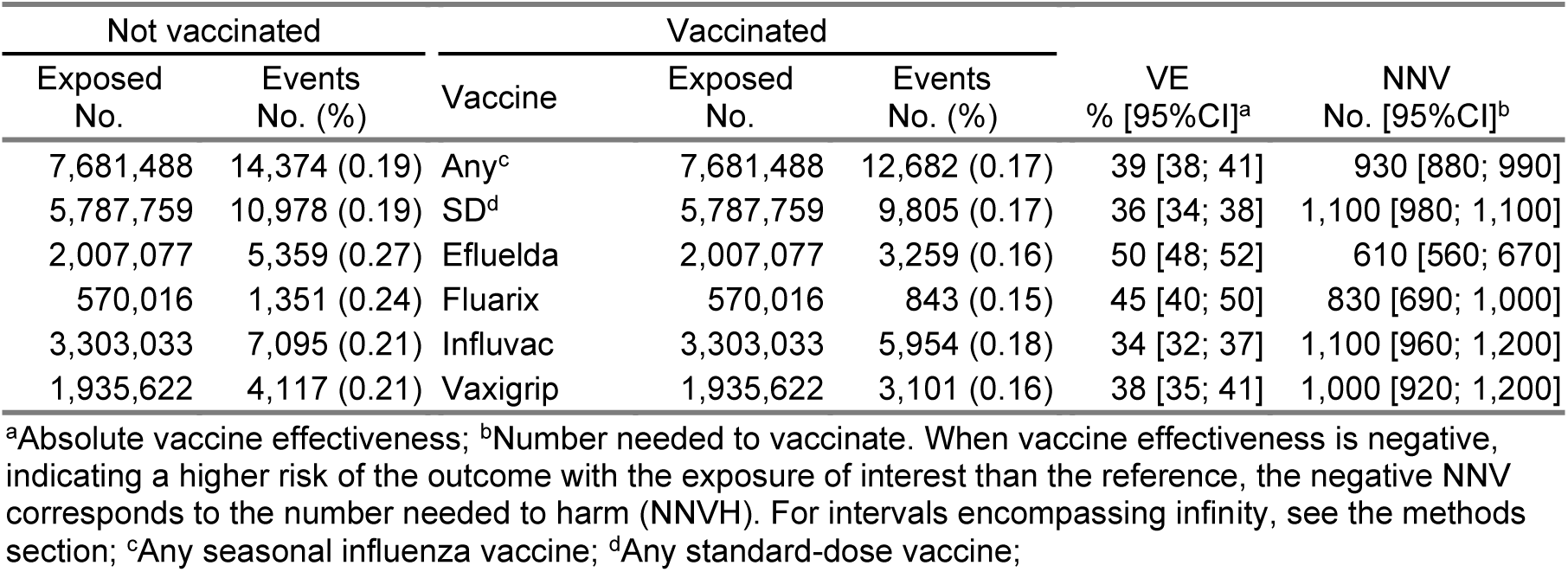
Absolute vaccine effectiveness against influenza-related hospitalization.

SD vaccines in terms of influenza-related hospitalization (50% vs 36%, Table 3). These VE estimates translate into NNV to avert one hospitalization of 610 (95%CI, 560 to 670) for the HD vaccine, and 1,100 (95%CI, 980 to 1,100) for any SD vaccine. However, this analysis does not show a significant difference in protection against death conferred by the HD vaccine compared with the other vaccines (eTable *8*). eFigures *4* and *5* show the cumulative incidence curves in unvaccinated and vaccinated groups for the secondary and negative outcomes, and the corresponding estimates of overall VE on all outcomes are shown in eTable *9*. Estimates regarding the risk of all cause in-hospital death, pneumonia, or of cardiac disorder suggest a protective effect of vaccination but one of a similar magnitude as that observed for the risk of traumatic injury, chosen as a negative control outcome. In contrast, vaccinated individuals were more likely to be hospitalized for cataract during the season than unvaccinated ones. The estimated VE on hospitalization for influenza by age class are shown for any vaccination vs no vaccination in Figure 2(B), and for the HD or any of the SD vaccines vs no vaccination in Figure 2(C). Absolute effectiveness tends to consistently decrease with age. Respective estimates of VE for HD or SD vaccines cannot be differentiated for the lowest age class, from 64 to 69 years. However, VE of the HD vaccine is greater than that of SD vaccines for the other age classes from 70 years and over.

Standard errors from the two-way cluster-robust analysis were similar to the primary estimates, indicating no material impact of within-subject correlation across weekly risk sets (eTable *10*).

## Discussion

This study evaluates the effectiveness in the elderly population of the various vaccines against the severe forms of seasonal influenza in a nationwide setting. We find a 39% lower risk of influenza-related hospitalization and 45% lower risk of death during those hospitalizations compared with no vaccination, in the target population of older adults. The high-dose vaccine was associated with greater VE than standard-dose vaccines against hospitalization (50% vs 36%), a difference most evident among adults 70 years and older, and with significantly greater rVE against hospitalization (27%) as well.

Our results of overall vaccine effectiveness align with estimates from analogous observational studies among people aged 65 and over available in France and Europe for the 2023-2024 season. Estimates of VE in France are based primarily on sentinel surveillance of biologically confirmed symptomatic infection^12–14^. In 2023–2024, VE against infection, as measured in community settings, was estimated at 42% (95%CI, 37 to 47)^13^. The European VEBIS study provides estimates of effectiveness in terms of hospitalisation for severe acute respiratory infection (SARI), but these did not include French data^15–17^. In 2023– 2024, the effectiveness of vaccines against SARI, as measured in hospitals at European level, was estimated at 36% (95%CI, 22 to 47)^15^. These French or European studies, all based on a test-negative design, do not provide estimates by vaccine. In a study carried out in Nordic countries, which was similar to ours for the 2024-2025 season, the high-dose vaccine was also found to be more effective at protecting against influenza-related hospitalization than standard-dose vaccines^6^. The authors report confidence intervals of absolute effectiveness from 38.1 to 88.7% for Efluelda, 23.7 to 63.6% for Vaxigrip and −7.8 to 69.1% for Influvac, that are consistent with but less precise than our corresponding estimates for the 2023-2024 season of 48 to 52% for Efluelda, 35 to 41% for Vaxigrip and 32 to 37% for Influvac. However, our estimate of rVE of HD vs SD vaccines for influenza-related hospitalization (27% [95%CI, 23% to 31%]) is in the lower range of the reported estimates from a meta analysis of randomized clinical trials (26.5 to 48.5%)^2^.

The strength of our study is the ability to provide precise estimates of both the relative and absolute vaccine effectiveness on a large population scale. For each objective, we used study designs that control for differences in the characteristics of vaccinated and unvaccinated population, or within vaccinated by different products. We benefitted from the flexible allocation of the four different vaccines in France that allowed to compare each product within all age brackets of the elderly population. We further derived the number needed to vaccinate for both absolute and relative comparisons across age groups in this population, translating our findings into actionable measures of the cases and deaths prevented by vaccination itself and by choosing one product over another — a level of granularity rarely achieved in other settings.

Using distinct designs for each objective reflected the different exposure contrasts involved in our context. Head-to-head comparisons could rely on a landmark design, as product exposure is stable once assigned, and because the landmark date preceded the epidemic, no outcome occurred before exposure was set. It is worth noting that in a season where the epidemic starts earlier relative to the typically stable timing of the vaccination campaign (mid-October in France), a substantial fraction of eventual vaccinees would still be unexposed when the epidemic begins, making a landmark design less suitable. Absolute effectiveness required a different approach, as the unvaccinated state is transient, with individuals becoming exposed throughout the season as events accrue. We addressed this with a sequential trial emulation. However, a sequential design would have been unnecessary for relative effectiveness, as product-specific exposure was already determined before the epidemic started, leaving no time-varying process to account for.

This study has some limitations. As an observational study, our study may have been affected by confounding that typically differentiates vaccinated and unvaccinated indidividuals. We adressed this potential confounding by matching and propensity score based weighting procedures, and also importantly ensure that the two groups were followed up at the same periods through the epidemic by the sequential trial method. Nevertheless, we observed evidence of residual confounding in the evaluation of negative control outcomes. While the hazard ratio for traumatic injury and cataract are reasonably close to 1 between vaccine recipients of different products, those outcomes are found to be more strongly associated with whether or not a person has been vaccinated, with associations going in both directions. Hospitalization for cataracts more likely reflects a greater tendency to seek medical care, whereas traumatic injuries are indicative of risk-taking or frailty. We would argue that this latter control outcome follows a pattern more closely aligned with what distinguishes vaccinated from unvaccinated individuals in terms of the risk of severe influenza. Thus, the impact of unmeasured confounding would more likely be an overestimation of VE. Our data source did not allow to individually characterise the viral subtype involved in the influenza-related hospitalization. In 2023-2024 viral circulation was dominated almost exclusively by influenza type A, with a larger share of subtype A(H1N1)pdm09 than A(H3N2)^8^. Acknowledging that VE might differ by subtype^13,15^, our findings apply to the 2023-2024 epidemic as a whole and might not be generalized to a different circulation context.

In conclusion, this nationwide cohort study indicate that influenza vaccines confer a moderate protection against severe outcomes in older adults. High-dose vaccination was associated with greater effectiveness than standard-dose vaccines, particularly among adults 70 years and older. These findings support product-level differentiation in vaccination strategy for this population.

## Data Availability

According to the principles of data protection and French regulations, the authors are not allowed to publicly release the data from the French National Health Data System (SNDS). However, any person or structure, public or private, for-profit or non-profit, can access SNDS data upon authorization from the French Data Protection Office (CNIL Commission Nationale de l'Informatique et des Libertés) to carry out a study, research, or an evaluation of public interest (https://www.snds.gouv.fr/SNDS/Processus-d-acces-aux-donnees and https://www.health-data-hub.fr/depot)

## Contributions

Stéphane Le Vu had full access to all of the data in the study and takes responsibility for the integrity of the data and the accuracy of the data analysis.

Concept and design: *Le Vu, Zureik*

Acquisition, analysis, or interpretation of data: *Le Vu, Gutiérrez-Melo, Cartailler*

Drafting of the manuscript: *Le Vu*

Critical review of the manuscript for important intellectual content: All authors.

Statistical analysis: *Le Vu*

Supervision: *Olié, Weill, Dray-Spira, Zureik*

## Data sharing statement

Data available: No Additional Information Explanation for why data not available: According to the principles of data protection and French regulations, the authors are not allowed to publicly release the data from the French National Health Data System (SNDS). However, any person or structure, public or private, for-profit or non-profit, can access SNDS data upon authorization from the French Data Protection Office (CNIL Commission Nationale de l’Informatique et des Libertés) to carry out a study, research, or an evaluation of public interest (https://www.snds.gouv.fr/SNDS/Processus-d-acces-aux-donnees and https://www.health-data-hub.fr/depot).

