## Supplement 1 for "Absolute and Relative Effectiveness of High-Dose vs Standard-Dose Influenza Vaccines Against Severe Outcomes in Older Adults"

**Supplementary material for the article 'Absolute and Relative Effectiveness of High-Dose vs Standard-Dose Influenza Vaccines Against Severe Outcomes in Older Adults' by Le Vu et al.**

2026-09-29

### eMethods

#### Data Source

We used data from the National Health Data System (Système National des Données de Santé - SNDS) which covers the entire population of France (67 million residents). Each person is anonymously identified by a unique, lifelong number. Since 2006, the SNDS has recorded all reimbursement data for both outpatient care (including drugs, imaging, and laboratory tests) and inpatient care (including diagnoses and procedures performed), as well as fully reimbursed health expenditures for patients with long-term diseases, such as cancer and diabetes. Our research group (Epi-Phare) has a regulatory permanent access to the data from the SNDS. This access is granted according to the French Decree No. 2016-1871 of December 26, 2016 and French law articles Art. R. 1461-13 and 14. According to the French law, informed consent and ethical approval from an institutional review board are not required when analysing deidentified existing healthcare data for research purposes. The SNDS has been extensively used to conduct real-life pharmacoepidemiological studies, notably on vaccines[1–5].

#### Covariates

Complementary solidarity health insurance (CSS) is a state public health insurance program that provides health coverage to individuals with low income. Comorbidities within 5 years included: history of diabetes, chronic respiratory disease, cardiovascular disease, cerebrovascular disease, dementia, immunodeficiency, chronic liver disease, cancer, alcohol dependence, tobacco dependence, obesity care, hospitalization for influenza, and hospitalization for COVID-19.

#### Confidence intervals of number needed to vaccinate (NNV)

When the 95% confidence interval for VE included the null, the corresponding NNV confidence interval was discontinuous and reported as its two bounds bridging infinity, expressed as the number needed to vaccinate to benefit (B) and the number needed to vaccinate to harm (H), as adapted from Altman[6]. In such case, an interval of the form [H n1;∞;B n2 ] indicates that, within the bounds of the 95% CI, vaccinating fewer than n1 persons is not expected to cause one additional case, and vaccinating fewer than n2 persons is not expected to avert one case.

### Supplementary tables

eTable 1: Hospital diagnoses and corresponding ICD-10 codes

| Hospital diagnosis | ICD-10 code(s) |
|---|---|
| Influenza | J09–J11 |
| Pneumonia | J12–J18 |
| Cardiovascular disorder | I20–I22, I24, I26, I30, I40, I46–I50, I60–I66, G45, G46 |
| Traumatic injury | S* |
| Cataract | H25 |

eTable 2: Age class distribution of vaccine recipients.

| | Age class No (%) | | | | |
|---|---|---|---|---|---|
| Vaccine | 64-69 | 70-74 | 75-79 | 80+ | Total |
| Efluelda | 314,849 (15) | 451,527 (22) | 447,444 (22) | 849,384 (41) | 2,063,204 (100) |
| Fluarix | 129,018 (23) | 142,643 (25) | 122,781 (22) | 175,717 (31) | 570,159 (100) |
| Influvac | 730,684 (22) | 825,165 (25) | 722,388 (22) | 1,032,841 (31) | 3,311,078 (100) |
| Vaxigrip | 470,477 (24) | 487,395 (25) | 414,586 (21) | 563,651 (29) | 1,936,109 (100) |

eTable 3: Characteristics of persons vaccinated at baseline with high-dose (HD) or standard-dose (SD) vaccines.

| Characteristics | Exposure status, No. (%) | | | |
|---|---|---|---|---|
| | Before matching | | | After matching |
| | HD (N=1,788,051) | SD (N=3,901,126) | Total (N=5,689,177) | SD (N=3,901,126) |
| **Male** | 784,527 (44) | 1,779,101 (46) | 2,563,628 (45) | 1,719,124 (44) |
| **Age class, y** | | | | |
| 64-69 | 270,861 (15) | 869,405 (22) | 1,140,266 (20) | 590,958 (15) |
| 70-74 | 389,488 (22) | 947,206 (24) | 1,336,694 (23) | 849,775 (22) |
| 75-79 | 387,915 (22) | 834,434 (21) | 1,222,349 (21) | 846,343 (22) |
| 80+ | 739,787 (41) | 1,250,081 (32) | 1,989,868 (35) | 1,614,049 (41) |
| **Age, y**[a] | | | | |
| Mean (sd) | 78 (8.2) | 76 (7.9) | 77 (8.1) | 78 (8.2) |
| Median (Q1;Q3) | 77 (72; 85) | 75 (70; 82) | 76 (71; 83) | 77 (72; 84) |
| **Health solidarity**[b] | 83,168 (4.7) | 184,891 (4.7) | 268,059 (4.7) | 190,299 (4.9) |
| **Nursing home**[c] | 144,212 (8.1) | 119,313 (3.1) | 263,525 (4.6) | 314,302 (8.1) |
| **Region** | | | | |
| Île-de-France | 270,395 (15) | 495,934 (13) | 766,329 (13) | 589,941 (15) |
| Auvergne-Rhône-Alpes | 201,296 (11) | 463,131 (12) | 664,427 (12) | 439,183 (11) |
| Nouvelle-Aquitaine | 185,008 (10) | 444,808 (11) | 629,816 (11) | 403,646 (10) |
| Occitanie | 147,516 (8.3) | 406,692 (10) | 554,208 (9.7) | 321,847 (8.3) |
| Hauts-de-France | 200,994 (11) | 318,964 (8.2) | 519,958 (9.1) | 438,524 (11) |
| Grand-Est | 197,113 (11) | 293,622 (7.5) | 490,735 (8.6) | 430,056 (11) |
| Provence-Alpes-Côte d'Azur | 102,834 (5.8) | 342,629 (8.8) | 445,463 (7.8) | 224,361 (5.8) |
| Pays de la Loire | 96,394 (5.4) | 277,089 (7.1) | 373,483 (6.6) | 210,310 (5.4) |
| Bretagne | 93,102 (5.2) | 266,302 (6.8) | 359,404 (6.3) | 203,128 (5.2) |
| Normandie | 136,781 (7.6) | 199,746 (5.1) | 336,527 (5.9) | 298,425 (7.6) |
| Bourgogne-Franche Comté | 59,983 (3.4) | 205,503 (5.3) | 265,486 (4.7) | 130,869 (3.4) |
| Centre-Val-de-Loire | 92,648 (5.2) | 167,883 (4.3) | 260,531 (4.6) | 202,137 (5.2) |
| Corse | 3,987 (0.22) | 18,823 (0.48) | 22,810 (0.40) | 8,699 (0.22) |
| **No. of care contacts**[d] | | | | |
| Mean (sd) | 8.6 (7.5) | 8.2 (7.2) | 8.3 (7.3) | 8.6 (7.8) |
| Median (Q1;Q3) | 7.0 (4.0; 12) | 7.0 (4.0; 11) | 7.0 (4.0; 11) | 7.0 (4.0; 12) |
| **Comorbidities/History** | | | | |
| Diabetes | 407,177 (23) | 849,592 (22) | 1,256,769 (22) | 893,245 (23) |
| Chronic respiratory disease | 235,600 (13) | 496,075 (13) | 731,675 (13) | 525,795 (13) |
| Cardiovascular disease | 347,218 (19) | 686,461 (18) | 1,033,679 (18) | 773,686 (20) |
| Cerebrovascular disease | 41,028 (2.3) | 73,225 (1.9) | 114,253 (2.0) | 94,539 (2.4) |
| Dementia | 119,002 (6.7) | 148,773 (3.8) | 267,775 (4.7) | 258,106 (6.6) |
| Immunodeficiency | 322,395 (18) | 667,042 (17) | 989,437 (17) | 706,412 (18) |
| Chronic liver disease | 21,736 (1.2) | 46,776 (1.2) | 68,512 (1.2) | 53,869 (1.4) |
| Cancer | 318,105 (18) | 656,582 (17) | 974,687 (17) | 695,295 (18) |
| Alcohol dependence | 41,528 (2.3) | 86,930 (2.2) | 128,458 (2.3) | 101,813 (2.6) |
| Tobacco dependence | 115,423 (6.5) | 261,257 (6.7) | 376,680 (6.6) | 267,762 (6.9) |
| Obesity care | 113,444 (6.3) | 234,405 (6.0) | 347,849 (6.1) | 267,238 (6.9) |
| History of hospitalisation for influenza | 8,641 (0.48) | 15,564 (0.40) | 24,205 (0.43) | 21,364 (0.55) |
| History of hospitalisation for COVID-19 | 51,307 (2.9) | 88,891 (2.3) | 140,198 (2.5) | 119,396 (3.1) |

[a]sd : standard deviation. Q1;Q3: 1st and 3d distribution quartiles; [b]Affiliation to complementary solidarity health insurance (CSS, state public health insurance program that provides health coverage to individuals with low income); [c]Nursing home residency: indidividuals residing in nursing homes whose medication costs are reimbursed through a lump-sum allocation are exluded; [d]Number of primary care contacts within 12 months;

eTable 4: Relative vaccine effectiveness against death during influenza-related hospitalization.

| Reference exposure | | | Exposure of interest | | | | |
|---|---|---|---|---|---|---|---|
| Vaccine | Exposed No. | Events No. (%) | Vaccine | Exposed No. | Events No. (%) | rVE % [95%CI][a] | NNV No. [95%CI][b] |
| SD[c] | 3,901,126 | 631 (0.016) | HD[d] | 1,788,051 | 301 (0.017) | 27 [14; 38] | 16,000 [9,600; 37,000] |
| Fluarix | 389,770 | 58 (0.015) | Efluelda | 1,788,051 | 301 (0.017) | 19 [-17; 43] | 26,000 [H 40,000; ∞; B 7,700] |
| Influvac | 2,327,657 | 373 (0.016) | Efluelda | 1,788,051 | 301 (0.017) | 26 [11; 39] | 17,000 [9,200; 49,000] |
| Vaxigrip | 1,183,674 | 200 (0.017) | Efluelda | 1,788,051 | 301 (0.017) | 24 [2.6; 40] | 19,000 [8,900; 220,000] |
| Influvac | 2,327,657 | 373 (0.016) | Fluarix | 389,770 | 58 (0.015) | 19 [-8.9; 40] | 28,000 [H 82,000; ∞; B 9,900] |
| Vaxigrip | 1,183,674 | 200 (0.017) | Fluarix | 389,770 | 58 (0.015) | 22 [-7.5; 43] | 24,000 [H 96,000; ∞; B 8,900] |
| Influvac | 2,327,657 | 373 (0.016) | Vaxigrip | 1,183,674 | 200 (0.017) | -16 [-41; 4.9] | -44,000 [H 20,000; ∞; B 110,000] |

[a]Relative vaccine effectiveness; [b]Number needed to vaccinate. When vaccine effectiveness is negative, indicating a higher risk of the outcome with the exposure of interest than the reference, the negative NNV corresponds to the number needed to harm (NNVH). For intervals encompassing infinity, see the methods section; [c]Any standard-dose vaccine; [d]High-dose vaccine (here Efluelda);

eTable 5: Relative vaccine effectiveness of high-dose (HD) vs standard-dose (SD) vaccines on all outcomes.

| Outcomes | SD[a] (N=3,901,126) Events No. (%) | HD[b] (N=1,788,051) Events No. (%) | HR[c] [95%CI] | rVE[d] [95%CI] |
|---|---|---|---|---|
| **Primary** | | | | |
| Influenza hosp. | 7,309 (0.19) | 2,987 (0.17) | 0.73 [0.69; 0.77] | 27 [23; 31] |
| All-cause death during influenza hosp. | 631 (0.016) | 301 (0.017) | 0.73 [0.62; 0.86] | 27 [14; 38] |
| **Secondary** | | | | |
| Pneumonia | 39,027 (1.0) | 22,181 (1.2) | 0.98 [0.96; 1.0] | 1.7 [-0.23; 3.6] |
| Cardiac disorder | 163,913 (4.2) | 85,751 (4.8) | 0.98 [0.97; 0.99] | 1.6 [0.70; 2.6] |
| All-cause death during any hosp. | 23,811 (0.61) | 14,229 (0.80) | 0.99 [0.96; 1.0] | 1.1 [-1.4; 3.6] |
| **Negative** | | | | |
| Traumatic injury | 62,031 (1.6) | 34,008 (1.9) | 1.0 [0.99; 1.0] | -0.62 [-2.2; 0.93] |
| Cataract | 71,590 (1.8) | 31,053 (1.7) | 0.97 [0.96; 0.98] | 3.1 [1.6; 4.5] |

[a]Any standard-dose vaccine; [b]High-dose vaccine (here Efluelda); [c]Hazard ratio; [d]Relative vaccine effectiveness;

eTable 6: Relative vaccine effectiveness of high-dose vaccine on influenza-related hospitalization, by age class.

| Age class, y | SD[a] | | HD[b] | | rVE | NNV |
|---|---|---|---|---|---|---|
| | Exposed No. | Events No. (%) | Exposed No. | Events No. (%) | % [95%CI][c] | No. [95%CI][d] |
| 64-69 | 869,405 | 605 (0.070) | 270,861 | 170 (0.063) | 25 [10; 38] | 4,700 [2,600; 14,000] |
| 70-74 | 947,206 | 987 (0.10) | 389,488 | 295 (0.076) | 33 [23; 42] | 2,700 [1,800; 4,500] |
| 75-79 | 834,434 | 1,334 (0.16) | 387,915 | 491 (0.13) | 26 [17; 34] | 2,200 [1,500; 3,900] |
| 80+ | 1,250,081 | 4,383 (0.35) | 739,787 | 2,031 (0.27) | 28 [23; 32] | 940 [770; 1,200] |

[a]Any standard-dose vaccine; [b]High-dose vaccine (here Efluelda); [c]Relative vaccine effectiveness; [d]Number needed to vaccinate. When vaccine effectiveness is negative, indicating a higher risk of the outcome with the exposure of interest than the reference, the negative NNV corresponds to the number needed to harm (NNVH). For intervals encompassing infinity, see the methods section;

eTable 7: Risk sets in sequential trial of overall vaccine effectiveness.

| Risk set | Day start | Unexposed No. | Exposed No. | Matched unexposed No. (%) | Matched exposed No. (%) |
|---|---|---|---|---|---|
| 1 | 0 | 8,329,022 | 5,589,955 | 5,392,670 (65) | 5,392,670 (96) |
| 2 | 2 | 7,619,700 | 706,655 | 706,652 (9.3) | 706,652 (100) |
| 3 | 9 | 7,087,411 | 526,443 | 526,443 (7.4) | 526,443 (100) |
| 4 | 16 | 6,684,616 | 397,249 | 397,249 (5.9) | 397,249 (100) |
| 5 | 23 | 6,398,645 | 280,447 | 280,447 (4.4) | 280,447 (100) |
| 6 | 30 | 6,245,977 | 146,933 | 146,933 (2.4) | 146,933 (100) |
| 7 | 37 | 6,186,890 | 53,665 | 53,665 (0.87) | 53,665 (100) |
| 8 | 44 | 6,125,073 | 56,280 | 56,280 (0.92) | 56,280 (100) |
| 9 | 51 | 6,067,696 | 51,549 | 51,549 (0.85) | 51,549 (100) |
| 10 | 58 | 6,034,087 | 27,865 | 27,865 (0.46) | 27,865 (100) |
| 11 | 65 | 6,009,583 | 19,119 | 19,119 (0.32) | 19,119 (100) |
| 12 | 72 | 5,991,698 | 12,949 | 12,949 (0.22) | 12,949 (100) |
| 13 | 79 | 5,982,263 | 5,020 | 5,020 (0.084) | 5,020 (100) |
| 14 | 86 | 5,975,979 | 2,232 | 2,232 (0.037) | 2,232 (100) |
| 15 | 93 | 5,970,967 | 1,354 | 1,354 (0.023) | 1,354 (100) |
| 16 | 100 | 5,966,350 | 985 | 985 (0.017) | 985 (100) |
| 17 | 107 | 5,962,861 | 39 | 39 (0.00065) | 39 (100) |
| 18 | 114 | 5,959,616 | 20 | 20 (0.00034) | 20 (100) |
| 19 | 121 | 5,956,428 | 17 | 17 (0.00029) | 17 (100) |
| Total | | 120,554,862 | 7,878,776 | 7,681,488 (6.4) | 7,681,488 (97) |

eTable 8: Absolute vaccine effectiveness against death during influenza-related hospitalization.

| Not vaccinated | | Vaccinated | | | | |
|---|---|---|---|---|---|---|
| Exposed No. | Events No. (%) | Vaccine | Exposed No. | Events No. (%) | VE % [95%CI][a] | NNV No. [95%CI][b] |
| 7,681,794 | 1,325 (0.017) | Any[c] | 7,681,794 | 1,096 (0.014) | 45 [40; 49] | 8,600 [7,200; 10,000] |
| 5,788,007 | 1,001 (0.017) | SD[d] | 5,788,007 | 831 (0.014) | 42 [36; 47] | 9,600 [7,800; 12,000] |
| 2,007,124 | 531 (0.026) | Efluelda | 2,007,124 | 322 (0.016) | 52 [44; 58] | 5,900 [4,500; 7,900] |
| 570,030 | 139 (0.024) | Fluarix | 570,030 | 74 (0.013) | 52 [36; 64] | 7,100 [4,300; 14,000] |
| 3,303,162 | 648 (0.020) | Influvac | 3,303,162 | 483 (0.015) | 44 [37; 50] | 8,800 [6,800; 12,000] |
| 1,935,726 | 397 (0.021) | Vaxigrip | 1,935,726 | 286 (0.015) | 43 [33; 51] | 9,000 [6,500; 14,000] |

[a]Absolute vaccine effectiveness; [b]Number needed to vaccinate. When vaccine effectiveness is negative, indicating a higher risk of the outcome with the exposure of interest than the reference, the negative NNV corresponds to the number needed to harm (NNVH). For intervals encompassing infinity, see the methods section; [c]Any seasonal influenza vaccine; [d]Any standard-dose vaccine;

eTable 9: Overall absolute effectiveness of any influenza vaccination on all outcomes.

| Outcomes | Not vaccinated (N=7,681,488) Events No. (%) | Vaccinated (N=7,681,488) Events No. (%) | HR[a] [95%CI] | VE[b] [95%CI] |
|---|---|---|---|---|
| **Primary** | | | | |
| Influenza-related hospitalisation | 14,374 (0.19) | 12,682 (0.17) | 0.61 [0.59; 0.62] | 39 [38; 41] |
| Death during influenza-related hospitalisation | 1,325 (0.017) | 1,096 (0.014) | 0.55 [0.51; 0.60] | 45 [40; 49] |
| **Secondary** | | | | |
| Pneumonia | 54,725 (0.71) | 73,483 (0.96) | 0.92 [0.91; 0.93] | 8.0 [7.0; 9.1] |
| Cardiac disorder | 218,467 (2.9) | 309,317 (4.0) | 0.98 [0.97; 0.98] | 2.4 [1.9; 3.0] |
| All-cause death during any hospitalisation | 41,890 (0.55) | 45,943 (0.60) | 0.73 [0.72; 0.74] | 27 [26; 28] |
| **Negative** | | | | |
| Traumatic injury | 91,272 (1.2) | 117,329 (1.5) | 0.93 [0.93; 0.94] | 6.6 [5.8; 7.4] |
| Cataract | 88,518 (1.2) | 134,977 (1.8) | 1.2 [1.2; 1.2] | -16 [-17; -15] |

[a]Hazard ratio; [b]Absolute vaccine effectiveness;

eTable 10: Hazard ratio estimates with cluster-robust standard errors under alternative clustering structures.

| variance_type | coef | se | coef_lo | coef_up | HR | HR_lo | HR_up |
|---|---|---|---|---|---|---|---|
| cluster: weekly risk set | -0.5005 | 0.0124 | -0.5248 | -0.4762 | 0.6062 | 0.5917 | 0.6211 |
| cluster: subject | -0.5005 | 0.0140 | -0.5279 | -0.4731 | 0.6062 | 0.5898 | 0.6231 |
| cluster: two-way (CGM) | -0.5005 | 0.0140 | -0.5279 | -0.4731 | 0.6062 | 0.5898 | 0.6231 |

Estimation of overall vaccine effectiveness against influenza hospitalization was used. Coefficients and hazard ratios are identical across rows; only standard errors and confidence intervals vary by clustering choice. Robust sandwich (Huber-White) standard errors were computed clustering score residuals by: week (weekly matched risk set, accounting for shared propensity-score models within each week), subject (individual, accounting for reuse of unexposed comparators across risk sets), or two-way (both jointly; Cameron, Gelbach & Miller, 2011). Columns: coef: log hazard ratio; se: cluster-robust standard error; coef_lo/_up: 95%CI on the log scale; HR: exp(coef); HR_lo/HR_up = 95%CI on the hazard ratio scale.

### Supplementary figures

eFigure 1: Covariates balance between high-dose and standard vaccines users before and after matching.

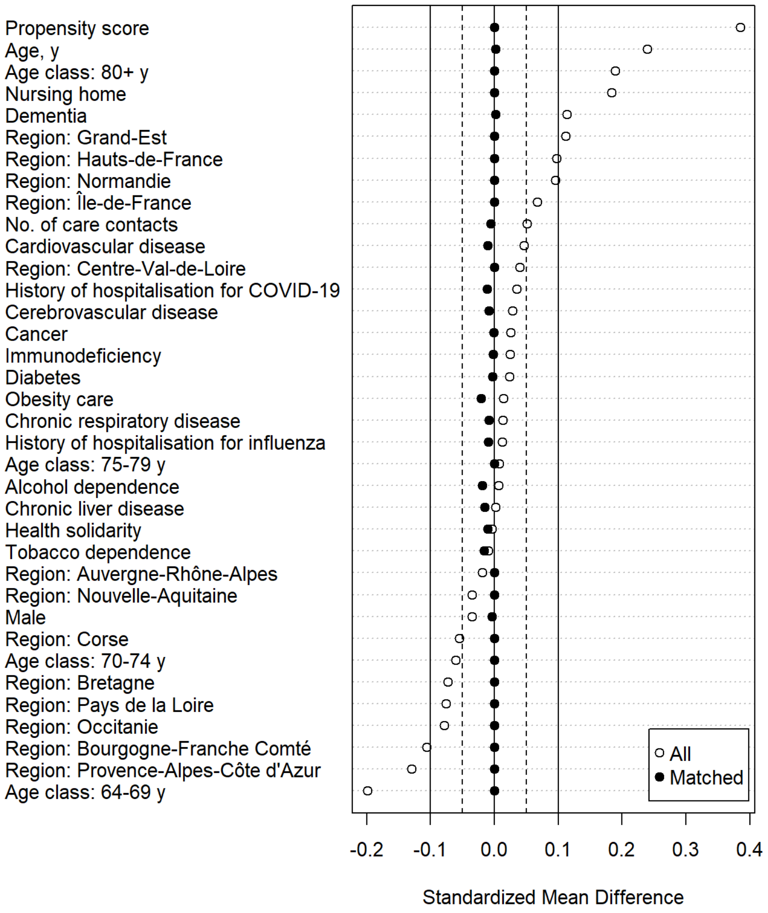

eFigure 2: Cumulative incidence curves for secondary outcomes - Landmark analysis.

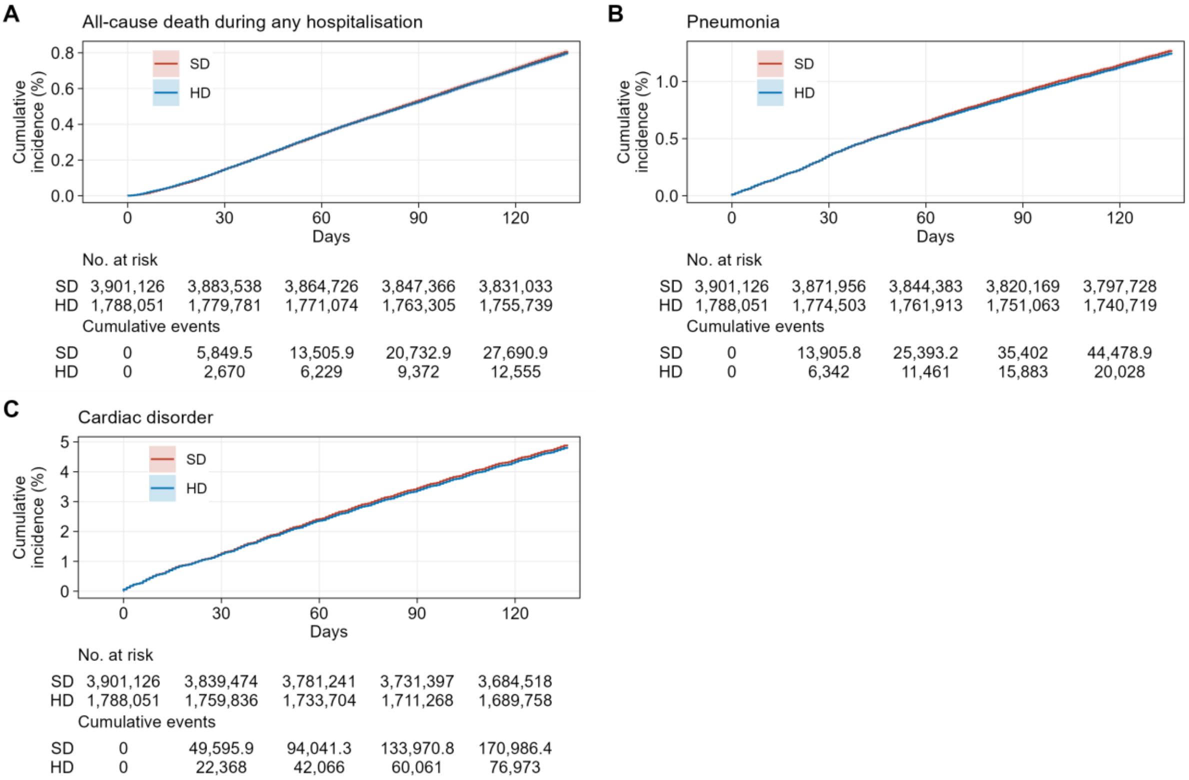


eFigure 3: Cumulative incidence curves for negative outcomes - Landmark analysis.

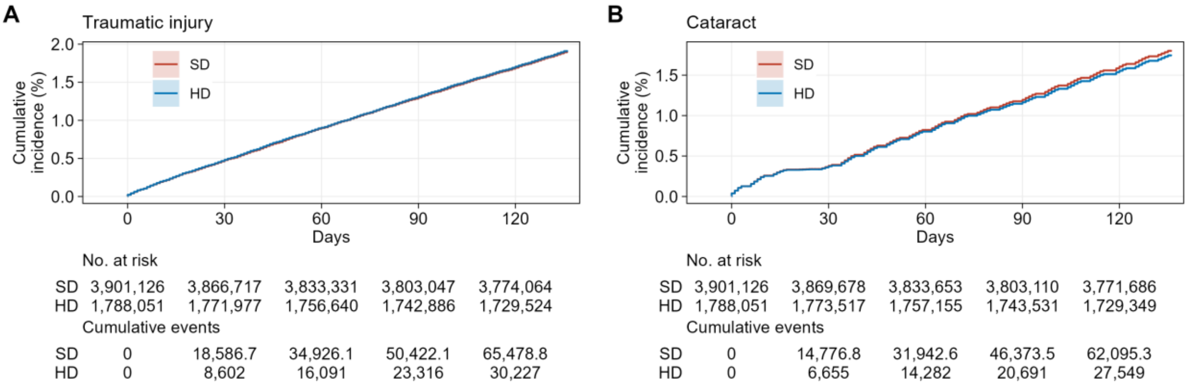

eFigure 4: Cumulative incidence curves for secondary outcomes - Sequential trial emulation.

**A** All-cause death during any hospitalisation

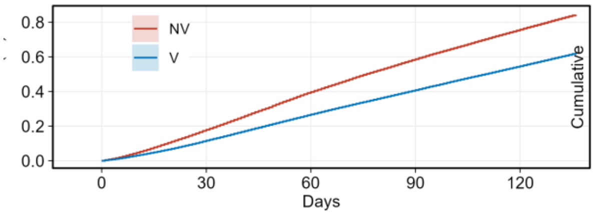


No. at risk

| | 0 | 30 | 60 | 90 | 120 |
|---|---|---|---|---|---|
| NV | 7,660,607 | 5,973,175 | 5,653,148 | 5,486,527 | 4,965,633 |
| V | 7,681,769 | 7,660,059 | 7,627,146 | 7,494,860 | 6,942,068 |

Cumulative events

| | 0 | 30 | 60 | 90 | 120 |
|---|---|---|---|---|---|
| NV | 0 | 11,615.9 | 24,210.1 | 34,845.6 | 43,897.5 |
| V | 0 | 8,952 | 20,506 | 31,192 | 41,274 |

**B** Pneumonia

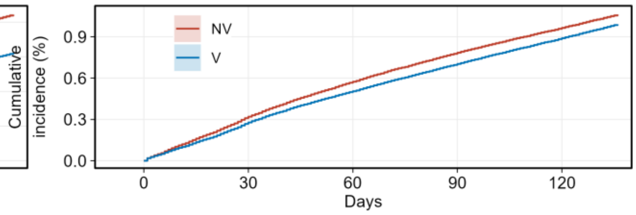


No. at risk

| | 0 | 30 | 60 | 90 | 120 |
|---|---|---|---|---|---|
| NV | 7,656,666 | 5,954,052 | 5,625,298 | 5,452,165 | 4,928,408 |
| V | 7,677,759 | 7,638,367 | 7,592,350 | 7,450,316 | 6,891,686 |

Cumulative events

| | 0 | 30 | 60 | 90 | 120 |
|---|---|---|---|---|---|
| NV | 0 | 21,051.7 | 35,705.1 | 47,461.9 | 56,972.7 |
| V | 0 | 21,307 | 38,649 | 53,677 | 67,262 |

**C** Cardiac disorder

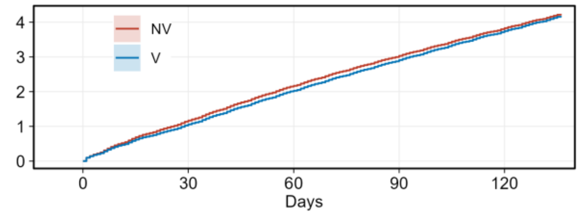


No. at risk

| | 0 | 30 | 60 | 90 | 120 |
|---|---|---|---|---|---|
| NV | 7,633,075 | 5,885,267 | 5,522,792 | 5,323,505 | 4,785,902 |
| V | 7,654,415 | 7,560,409 | 7,462,171 | 7,278,135 | 6,693,050 |

Cumulative events

| | 0 | 30 | 60 | 90 | 120 |
|---|---|---|---|---|---|
| NV | 0 | 77,675.1 | 135,201.1 | 183,366.8 | 225,460.4 |
| V | 0 | 80,896 | 154,785 | 221,561 | 282,381 |

eFigure 5: Cumulative incidence curves for negative outcomes - Sequential trial emulation.

**A** Traumatic injury

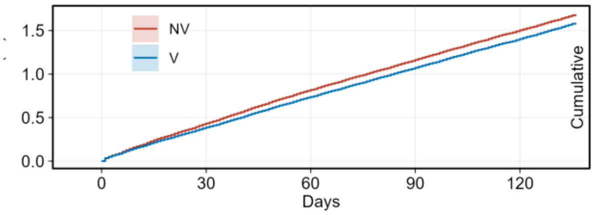


No. at risk

| | 0 | 30 | 60 | 90 | 120 |
|---|---|---|---|---|---|
| NV | 7,646,960 | 5,936,409 | 5,599,824 | 5,419,725 | 4,890,893 |
| V | 7,668,117 | 7,619,563 | 7,563,032 | 7,411,114 | 6,845,230 |

Cumulative events

| | 0 | 30 | 60 | 90 | 120 |
|---|---|---|---|---|---|
| NV | 0 | 28,734.2 | 50,930.6 | 70,195.2 | 88,096.3 |
| V | 0 | 29,692 | 56,488 | 81,816 | 105,886 |

**B** Cataract

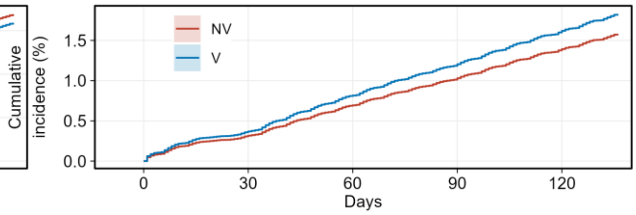


No. at risk

| | 0 | 30 | 60 | 90 | 120 |
|---|---|---|---|---|---|
| NV | 7,653,200 | 5,949,393 | 5,610,443 | 5,428,506 | 4,896,889 |
| V | 7,674,337 | 7,625,788 | 7,559,401 | 7,401,426 | 6,828,714 |

Cumulative events

| | 0 | 30 | 60 | 90 | 120 |
|---|---|---|---|---|---|
| NV | 0 | 21,682.3 | 43,409.6 | 62,224.7 | 81,358.8 |
| V | 0 | 28,263 | 62,360 | 91,872 | 122,068 |
